# Detecting microbiome species unique or enriched in 20+ cancer types and building cancer microbiome heterogeneity networks

**DOI:** 10.1101/2024.03.23.24304768

**Authors:** Zhanshan (Sam) Ma, Lianwei Li, Jiandong Mei

## Abstract

It is postulated that tumor tissue microbiome is one of the enabling characteristics that either promote or suppress cancer cells and tumors to acquire certain hallmarks (functional traits) of cancers, which highlights their critical importance to carcinogenesis, cancer progression and therapy responses. However, characterizing the tumor microbiomes is extremely challenging because of their low biomass and severe difficulties in controlling laboratory-borne contaminants, which is further aggravated by lack of comprehensively effective computational approaches to identify unique or enriched microbial species associated with cancers. Here we take advantages of two recent computational advances, one by Poore *et al* (2020, *Nature*) that computationally generated the microbiome datasets of 33 cancer types [of 10481 patients, including primary tumor (PT), solid normal tissue (NT), and blood samples] from whole-genome and whole-transcriptome data deposited in “The Cancer Genome Atlas” (TCGA), another termed “specificity diversity framework” (SDF) developed recently by Ma (2023). By reanalyzing Poore’s datasets with the SDF framework, further augmented with complex network analysis, we produced the following catalogues of microbial species (archaea, bacteria and viruses) with statistical rigor including unique species (USs) and enriched species (ESs) in PT, NT, or blood tissues. We further reconstructed species specificity network (SSN) and cancer microbiome heterogeneity network (CHN) to identify core/periphery network structures, from which we gain insights on the codependency of microbial species distribution on landscape of cancer types, which seems to suggest that the codependency appears to be universal across all cancer types.

## Introduction

It was already known that human tumor tissues contain bacteria more than a century ago; however, characterization and understanding of the tumor microbiomes are still very limited due to their relatively low biomass and severe experimental obstacles in controlling laboratory-borne contaminations (Nejman *et al*. 2020, Poore *et al*. 2020, Fu *et al*. 2022). The intratumor microbes are primarily intracellular and are distributed in both cancer and immune cells, but it is still not established whether there is a causal relationship between the intratumor bacteria and the development of cancer or whether their existence simply reflects infections of established tumors or whether they may come from adjacent normal tissues (Nejman *et al*. 2020), although progresses have been made recently (*e.g.,* Fu *et al*. 2022). Despite these uncertainties, some consensuses have emerged from recent laboratory and computational investigations. Nejman *et al* (2020) demonstrated that human tumor microbiome consists of tumor type-specific intracellular bacteria, and some species are enriched in certain tumor types. They further revealed that some metabolic functions encoded by intratumor bacteria are associated with disease features of certain tumor types (Nejman *et al*. 2020).

While Nejman *et al*. (2020) study relied on wet-lab experimental and amplicon sequencing procedures to characterize the PT (primary tumor) and NT (adjacent normal tissue) microbiomes from 7 cancer types, Poore *et al*. (2020) computational study reexamined the microbial reads of 33 cancer types (including a total of 18116 samples collected from 10481 patients) deposited in the TCGA (The Cancer Genome Atlas) database. Through highly sophisticated and finely designed bioinformatics pipelines, they were able to extract the microbial OTU tables (including archaea, bacteria and viruses) at species level for the PT, NT and blood samples from the whole-genome sequencing (WGS, *n*=4381) and whole-transcriptome sequencing (RNA-Seq, *n*=13285). More importantly, they applied machine learning algorithms, specifically the stochastic gradient-boosting machine (GBM) learning models to investigate the feasibility to use microbial signatures for cancer diagnoses and monitoring. Their machine learning analyses demonstrated unique microbial signatures in tissue and blood within and between most major types of cancer.

To the best of our knowledge, Poore *et al*. (2020) and Nejman *et al*. (2020) still represent the most comprehensive cancer tissue microbiome datasets from computational and experimental studies, respectively, to date besides some in-depth studies on individual cancer type such as breast tumor (Fu *et al*. 2022). In a pair of computational studies [this article and Ma (2023)], we take advantages of Poore *et al*. (2020) and Nejman *et al*. (2020) rich datasets of cancer microbiomes. In the first article (Ma 2023), we propose, test and apply a new framework, termed “specificity diversity framework” (SDF), for detecting the treatment effects (*e.g*., the differences between the healthy and diseased microbiome samples such as the PT and NT) at both OTU and assemblage or community levels. We developed and tested our framework with Nejman *et al*. (2020) dataset in previous article (Ma 2023). In the present article, our focus is on the practical application of the SDF—producing the catalogues of unique and enriched microbial species in 33 cancer types by reanalyzing Poore *et al*. (2020) and further performing specificity network analysis to quantitatively characterize the findings from applying the SDF. As a side note, the exceptional breadth of Poore *et al*. (2020) datasets hinted us that separate reporting of SDF methodology (Ma 2023) and this application here aimed to characterize cancer-associated microbiomes comprehensively should be a better option.

It should be noted that most existing studies on the microbiome-cancer relationships have been focused on the symbiotic microbes inhabited in or on the so-termed “barrier tissue” of the body exposed to the environment—the epidermis and the internal mucosa (particularly, gastrointestinal tract, lung, breast, urogenital system, and skin) (*e.g*., Zackular et al. 2014, Tsay *et al*. 2020, Hanahan 2022), which were the initial target of the NIH’s human microbiome project (iHMP 2019). Many microbiome datasets have been collected from those HMP-kind studies related to cancers, but they are different from the two cancer-tissue microbiome studies (Nejman *et al*. 2020, Poore *et al*. 2020) discussed previously, and we ignore those non-tumor-tissue datasets totally in the present article. Nevertheless, it is speculated that the mechanistic relationships between cancers and microbiomes involving both tumor-tissue and non-tumor-tissue (barrier tissue) microbiomes are likely similar (Hanahan 2022).

According to Hanahan’s (2000, 2011, 2022) conceptualization of cancer hallmarks, there are hallmarks of cancer that are generally applicable to all cancer types. Initially, the so-termed hallmarks of cancer refer to a set of eight functional capacities acquired by human cells of a potential cancer patient, including inducing or accessing vasculature, activating invasion & metastasis, enabling replicative immortality, avoiding immune destruction, evading growth suppressors, sustaining proliferative signaling, deregulating cellular metabolism, and resisting cell death (Hanahan 2000). However, these hallmarks *per se* cannot explain the complexities of cancer pathogenesis or the precise molecular and cellular mechanisms of malignant development progression. For this, Hanahan (2022) proposed the so-termed “enabling characteristics” that provide tools (means) for cancer cells and tumors to adopt those functional traits (capacities or the hallmarks). Initially, two enabling characteristics Hanahan (2000, 2022) proposed are genome instability & mutation, and tumor-promoting inflammation. Later, Hanahan (2022) added several prospective new hallmarks and enabling characteristics which may in due course be established as core constituents of the cancer hallmark scheme. These newly identified hallmark or enabling characteristics include tumor microenvironment (TME), unlocking phenotypic plasticity, non-mutational epigenetic reprogramming, polymorphic microbiomes and senescent cells. According to Hanahan (2022), the TME generally can be defined as “to be composed of heterogeneous and interactive populations of cancer cells and cancer stem cells along with a multiplicity of recruited stromal cell types”, which are believed to play an integral role in n tumorigenesis and malignant progression. Some scholars actually classified tumor tissue microbiome as part of the TMP given that they may live inside cancer and immune cells (Ma *et al*. 2021).

It has been revealed that the gut microbiome is implicated in the susceptibility, development and pathogenesis of colon cancer (Hanahan 2000, 2011, 2022; Fulbright *et al*. 2017; Tsay *et al*. 2020). The results from animal models confirmed the existence of cancer-protective and tumor-promoting microbiomes, involving *specific* bacterial species that can modulate the incidence and pathogenesis of colon cancers. Gut microbiomes are involved in the modulation of the adaptive and innate immune systems, through multifarious routes such as producing immunomodulatory factors. For example, *particular* and *variable* components of the gut microbiome may systemically modulate the activity of the adaptative immune system, either enhancing antitumoral immune responses induced by immune checkpoint blockade, or rather prompting systemic or local (intertumoral) immunosuppression. The systemically and indirectly modulating of tumor immunobiology is above and beyond immune responses due to direct physical interactions between bacteria and the immune system. Furthermore, the elicitation by gut microbiome of the expression of immunomodulatory chemokines and cytokines that enter the systemic circulation may also influence the cancer progression and response to therapy in other organs of the body such as the liver. Compared with the gut microbiome, the roles of the microbiomes inhabited in other host sites such as lung (*e.g*., Tsay *et al*. 2020) involving in homeostasis, aging and cancer have also been actively studied but with limited breakthroughs. Non-causal and association studies are revealing evidence that local tumor-antagonizing/protective *versus* tumor-promoting tissue microbiomes may modulate susceptibility and pathogenesis to cancers arising in the local organs. An open question is that whether microbiomes in different tissues have the capacity to contribute or interfere with the acquisition of other hallmark capacities beyond immunomodulation and genome mutation, consequently impacting cancer development and progression. There is emerging evidence supporting the possibilities that the human microbiomes are a distinctive enabling characteristic for the acquisition of cancer hallmark capabilities, beyond intersecting with and complementing those of genome instability and mutation as well as tumor-promoting inflammation, the two primary enabling characteristics initially identified by Hanahan (2000, 2011, 2022), also see Fulbright et al. (2017).

From above introductory summary on the cancer-microbiome relationships, it is clear that identifying *specific* microbial species in the cancer-associated microbiomes should be among the first steps in mechanistic investigations on the “enabling characteristics roles” possibly played by the human microbiomes, which provide means by which cancer cells and tumors can adopt hallmark traits. Obviously, much of the essential mechanistic results ultimately are linked to specific microbial species (or OTUs), rather than the whole microbiota, although both species level and microbiota (community) level insights are important. Despite enormous efforts both experimentally and computationally, the methods for detecting the difference between cancer and cancer microbiomes are still highly sought, which in turn have stalled the detection of the differences. In the present study, we take advantages of both a recent methodology advance termed SDF (specificity diversity framework) (Ma 2023) and the extensive cancer-tissue microbiome datasets by Poore *et al*. (2020), and we produce catalogues of unique (exclusive) species and enriched species in the PT, NT and blood samples, respectively. We further perform species specificity network (SSN) analysis to seek insights on the codependency of microbial species distribution on landscape of cancer types.

## Material and Methods

### Datasets of Cancer Microbiomes

Poore *et al*. (2020), with their highly sophisticated and powerful bioinformation pipeline, generated the microbial (bacteria, archaea and viruses) reads of 33 cancer types from the whole-genome sequencing (WGS) and whole transcriptome sequencing (RNA-Seq) data deposited in TCGA (The Cancer Genome Atlas) database, including a total of 18116 samples collected from 10481 patients. We excluded the datasets of 10 cancer types that do not have normal tissue (NT) controls, and used the datasets of remaining 23 cancer types, which have primary tumor (PT), solid normal tissue (NT) and blood (B) samples for our specificity diversity framework (SDF) analysis (briefly introduced in the next subsection). Table 1 briefly summarized the dataset information relevant to our SDF analysis.

**Table 1.** Brief description of the tumor-tissue microbiome datasets recalculated by Poore *et al*. (2020) from TCGA (The Tumor Genome Atlas) database for 23 cancer diseases (types)

| Aspects of Datasets | Descriptions |
| --- | --- |
| Data Sources | http://cancermicrobiome.ucsd.edu/CancerMicrobiome_DataBrowser/<br>https://gdc.cancer.gov/ |
| Software Packages | Kraken, GBM (stochastic Gradient-boosting Machine learning) and other software packages by Knight Lab of UC San Diego, Moores Cancer Center. https://knightlab.ucsd.edu/ |
| 33 Cancer Diseases (Types) belong to six Human Organ System | **Digestive system** (Cholangiocarcinoma, Colon Adenocarcinoma, Esophageal Carcinoma, Head and Neck Squamous Cell Carcinoma, Liver Hepatocellular Carcinoma, Pancreatic Adenocarcinoma, Rectum Adenocarcinoma, Stomach Adenocarcinoma) |
|  | **Urinary system** (Adrenocortical Carcinoma, Bladder Urothelial Carcinoma, Kidney Chromophobe, Kidney Renal Clear Cell Carcinoma, Kidney Renal Papillary Cell Carcinoma, Pheochromocytoma and Paraganglioma), Respiratory system (Head and Neck Squamous Cell Carcinoma, Lung Adenocarcinoma, Lung Squamous Cell Carcinoma) |
|  | **Reproductive system (M)** (Prostate Adenocarcinoma, Testicular Germ Cell Tumors) |
|  | **Reproductive system (F)** (Cervical Squamous Cell Carcinoma and Endocervical, Adenocarcinoma, Ovarian Serous Cystadenocarcinoma, Uterine Carcinosarcoma, Uterine Corpus Endometrial Carcinoma) |
|  | **Central nervous system** (Brain Lower Grade Glioma, Glioblastoma Multiforme) |
|  | **Others** (Breast Invasive Carcinoma, Lymphoid Neoplasm Diffuse Large B-cell Lymphoma, Mesothelioma, Sarcoma, Skin Cutaneous Melanoma, Thyroid Carcinoma, Thymoma, Uveal Melanoma). |
| Sampling sites of the cancer-associated microbiome | Primary tumor (PT), Solid normal tissue (NT), Blood derived normal (B)<br>(WGS: 1827 B Samples + 2117 PT Samples + 229 NT Samples; RNA-Seq: 11741 PT Samples + 641 NT Samples) |
| Reads Statistics Per Sample | Range=16,374—134,372,465; Mean=1,740,242; Standard Error=26267; Medium=103,194 |
| Sequencing Protocol | WGS (whole-genome sequencing: 4173 Samples); Whole transcriptome sequencing (RNA-Seq: 12382 Samples) |
| Microbial Taxa | Archaea, Bacteria, Viruses and their combined (Total); Microbial species abundance tables for these taxa are available at the site of *data sources* listed previously. |

### Categorizing microbial species with specificity diversity framework (SDF)

Inspired by Mariadassou *et al*. (2015) *species specificity* (SS) concept and metric and its application such as Ma & Li (2017), Ma (2023) introduced the concept and metric of *specificity diversity* (SD) and further developed a two-stage statistical test algorithm based on the two metrics and the principles of permutation tests (*e.g*., Pesarin & Salmaso 2010), collectively termed specificity diversity framework (SDF). The concept of *specificity diversity* can be considered as a new type of biodiversity and can be measured similarly as species (abundance) diversity or gene diversity with entropy. The advantage of entropy over other statistical moments such as mean or variance is that entropy is more effective in measuring uncertainty, similarity/dissimilarity, which may significantly supplement the discerning power of the original species specificity by Mariadassou *et al*. (2015). In fact, we use the Hill numbers (a function of Renyi’s entropy) that has achieved wide success in measuring species (abundance) diversity and other forms of biodiversity including gene and metagenome diversity (Chao *et al*. 2012, 2014, Ma & Li 2017, 2018, Ma *et al*. 2019). The permutation test had also been used in the original specificity test for testing a key hypothesis that specificity and local abundance is positively correlated in microbial ecosystems (across a wide array of habitats, sampling conditions and sequencing depths) in general (Mariadassou *et al*. 2015, Ma & Li 2017). This hypothesis is of critical significance for understanding microbial biogeography, and it also give us great confidence in using microbial abundance data broadly in this study since Mariadassou *et al*. (2015) study found that the pattern of specificity-abundance correlation is rather robust and is little influenced by sampling conditions and sequencing protocols (including sequencing depth). The use of permutation tests in our SDF was designed to detect the differences between treatments (*e.g*., PT and NT) in either species specificity (SS) or specificity diversity (SD) and is more similar to their usage in Ma *et al*. (2019) for shared species analysis in microbiomes.

The SDF was developed with two interdependent objectives: (*i*) identify species (OTUs) that are unique (exclusive) or enriched (upregulated) in a treatment (such as PT or NT), and therefore being able to compile lists of unique species (USs) and enriched species (ESs) in each treatment; (*ii*) detect the holistic difference between two assemblages (*e.g*., groups of unique or enriched species) or between two treatments (*e.g*., PT and NT).

Fig 1 illustrated the components of SDF (Ma 2023), tailored for the reanalysis of cancer microbiome datasets of Poore *et al*. (2020) with our objective to categorizing cancer-tissue microbial species as unique and/or enriched species associated with PT or NT, respectively. For detailed definitions, formulae, algorithms and computational codes of the SDF framework, readers are referred to Ma (2023). Here, we reiterate selected key concepts and principles of the SDF illustrated in Fig 1 and detailed in Ma (2023) for the practical purpose to present and interpret our results in the following sections. We summarize the computational procedures, with reference to the diagram illustrated with Fig 1, as the following three major blocks (steps).

**Fig 1.**
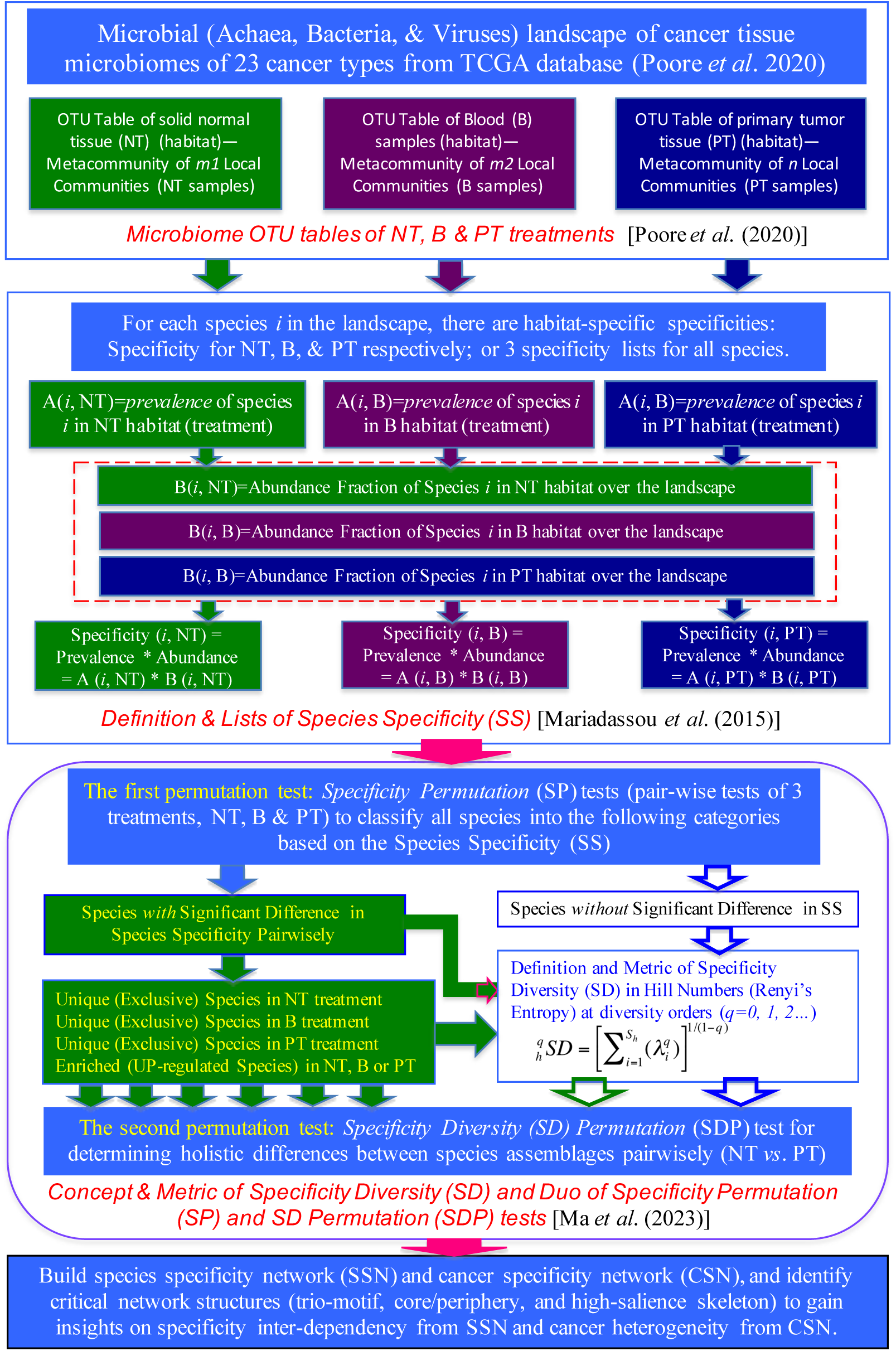
Diagraming the “specificity diversity framework” for cataloguing unique/enriches species in the cancer tissue microbiomes of 23 cancer types and for testing the specificity diversity, as well as their complex network analyses.

The top block in the diagram (Fig 1) simply shows the OTU tables of three treatments (*i.e.,* PT, NT and B) of the tissue microbiome samples of 23 cancer types originally published by Poore *et al*. (2020). Little special explanation is needed here about those OTU tables since we take advantages of the enormous quality control efforts taken in the bioinformatics pipelines of Poore *et al*. (2020) when they generated the datasets computationally from the TCGA database. An additional advantage of Poore *et al*. (2020) dataset is the virome datasets, which are still rare in current cancer microbiome studies. Overall, the top block represents the “input data” to our study for categorizing the microbial species as groups (assemblages) of unique, enriched, impoverished, or unchanged in terms of the species specificity (SS), and for detecting the differences between assemblages holistically in terms of the specificity diversity (SD).

The middle block (Fig 1) shows the SS of Mariadassou *et al*. (2015), which was apparently inspired by the specialist-generalist paradigm by Dufrene & Legendre (1997), and was designed to characterize the differences among habitats in terms of the species abundance distributions, which theoretically can fully determines the species composition and diversity of communities. As illustrated in Fig 1, the SS, specificity of a *species* is defined as product of its local prevalence (percentage of samples with the species in the local habitat or treatment such as PT) and its abundance share (fraction of its abundance across all habitats including PT, NT and B globally). Therefore, species specificity is both species and habitat (treatment) specific, and each species in the cancer microbiome of each cancer type may have up to 3 specificity values (in PT, NT, and B respectively). That is, there are up to three specificity lists for each cancer type. The SS takes a value of [0, 1] and is hence termed as *specificity continuum*, with *0* being absent locally and *1* being present exclusively locally.

The bottom block in the diagram (Fig 1) fulfills the objectives of this study by processing the possibly three *specificity lists* (for PT, NT and B) computed by the specificity definition (the middle block) for each of the 23 cancer types. Methodologically, the block includes the concept of specificity diversity (SD) and its metric, as well as a duo of permutation tests, the specificity permutation (SP) test and specificity-diversity permutation (SDP) test.

The SP test classifies all species in one treatment pair (*e.g*., PT & NT) into unique (exclusive), enriched (up-regulated), impoverished (down-regulated) or unchanged (indifferent) categories (groups or assemblages of species) exclusively. Note that enriched species (up-regulated) assemblage in the PT treatment corresponds improvised (down-regulated) species assemblage in the NT treatment, assuming that the pair-wise SP test is performed for the pair of the PT and NT treatments. Similarly, unique species in the PT correspond to absent species in the NT. Therefore, for a pairwise SP test (*e.g.,* PT and NT), we only need to produce four species lists reciprocally, unique species (US) in PT, unique species (US) species in NT, enriched species (ES) in PT, and enriched species (ES) in NT, as well as a fifth group in which the specificity is *indifferent* between PT and NT (no difference from random variation). The SP test, following the principle of standard permutation test, is performed with 1000 times of re-sampling, and a *P*-value=0.05 is adopted as threshold for determining the significance of differences. If a species’ specificity value is not significantly different from zero (SS=0) (*P*-value≤0.05), then the species is absent in the local habitat (say, the PT treatment), and it is considered as a unique species in the reciprocal treatment (NT). Similarly, if a species’ specificity value is significantly higher in the PT treatment than that in the NT treatment, as determined by the permutation test, then the species is classified as enriched species in the PT treatment, or impoverished species in the NT treatment.

The SDP test, again based on the principle of standard permutation test with the same operational setting (1000 times of re-sampling and threshold *P*-value=0.05), is designed to determine the difference between two assemblages (such as the assemblage of the enriched species in PT and the assemblage of the enriched species in the counterpart NT) in their specificity diversity values at different diversity order (*q*=0, 1, 2, 3, 4). Besides testing the four pairs of species assemblages (*e.g*., unique in PT or NT, enriched in PT or NT), we also tested the assemblages of “all species with significant difference in SS values”, as well as “all species” (without considering the SS difference) communities. Note that when diversity order *q*=0, the specificity diversity (the Hill number for *q*=0) defaults to the *species richness* in traditional species diversity because specificity value does not weigh in the calculation of SD. When the diversity order *q*=1, the specificity diversity is weighted in proportion to the relative level of species specificity, and is an exponential function of Shannon entropy. The SD value for *q*=1 then represents the equivalent number of species with *typical* specificity level. When *q*>1, the SD is weighted in favor for species with higher specificity level, which generally are locally more prevalent or more abundant species, or the both.

### Species specificity network (SSN) and cancer microbiome heterogeneity network (CHN)

Note the apparent similarity between *species specificity lists* (distribution) (SSD) and traditional species abundance distributions (SAD) in terms of their format: both species specificity lists and species abundance distributions are in the form of matrix. Therefore, complex network analysis can be applied to investigate the multivariate relationships among microbial species in terms of their correlation relationships measured in species specificity across cancer types, we term this type of complex network (with species or OTU as network nodes) we build, *species specificity network* (SSN). Alternatively, one can also build network of cancer types (as network nodes) by leveraging their correlation relationships in terms of the microbial species specificity across microbial species, we term this latter type of complex network, *cancer microbiome heterogeneity network* (CHN). With the SSN, the focus is on the interactions between microbial species, more specifically the *specificity codependency* between microbial species across cancer types. With the CHN network, the focus is on the similarity or differences between cancer types in terms of the microbial specificity codependency; we term this type of similarity or difference between cancers as a kind of cancer *homogeneity* or *heterogeneity*. Of course, the term heterogeneity has been widely used in oncology, and we should emphasize that the *heterogeneity* concept here emphasizes the *heterogeneity of specificity co-dependency patterns* among difference cancer types. In fact, the heterogeneity here is similar to previously introduced specificity diversity (SD), but in a network setting.

We followed the same computational algorithms and procedures previously used for microbial species correlation networks such as Ma & Ellison (2018, 2019, 2021), Ma (2020). Specifically, we build and analyze the core/periphery network (CPN) (Borgatti & Everett 2000, Csermely *et al*. 2013, Gallagher *et al*. 2021) with species specificity lists (matrices) (Ma & Ellison 2019, 2021; Ma 2020). The core/periphery network distinguishes all network nodes as core nodes and periphery nodes: with the former consisting of densely interconnected nodes, with the latter nodes sparsely connected to some core nodes. In an idealized core/periphery structures, there are no links within periphery nodes. We postulate that network structures such as core/periphery structures should be different between the specificity networks of tumor and normal tissue microbiomes.

Both the SSN and CHN previously designed appear to be similar to the *species cooccurrence* networks that have been extensively studied in the last decade or so. The difference is that specificity is dependent on both prevalence and global (meta-community or landscape) abundance fraction, while traditional species abundance is a local community property. In other words, the perspective of SSN and CHN are from metacommunity/landscape scales, while the perspective of traditional species cooccurrence networks is from community/metacommunity scales.

## Results

### Identify and cataloguing unique and enriched species lists for each cancer type

For each of the 23 cancer types, we first computed their specificity value lists for all treatments (PT, NT, blood) of each cancer type dataset (Table S1). The higher specificity values represent either higher local prevalence, high local abundance share (out of global or all treatments of the cancer type), or the both, and vice versa. Table S1A-S1H (8 sheets) exhibited the specificity values for: “archaea from RNA-Seq”, “bacteria from RNA-Seq”, “viruses from RNA-Seq”, “whole (all three taxa pooled) from RNA-seq”, “archaea from WGS”, “bacteria from WGS”, “viruses from WGS”, and “whole (all three taxa pooled) from WGS”. In each of the 8 sheets, the specificity lists for each cancer type were listed one type after another. For each cancer type, two columns of SS values, one for the NT samples (treatment) or blood (B) samples (treatment) representing the healthy (*H*) treatment, and another for the primary tumor (PT) samples (treatment) representing the diseased (*D*) treatment. The NT and blood treatments are traditionally considered as *healthy* treatments, and the PT is considered as *diseased* treatment, although all three kinds of samples are taken from cancer patient (Nejman *et al*. 2020, Poore *et al*. 2020). We followed this convention in this study. In the majority (21 out of 23) of cancer types, we have two treatments only: either NT or blood as the healthy, and PT as diseased. However, in two of the cancer types (datasets), we have three treatments, *i.e*., two healthy controls (NT & Blood) and diseased (PT). In the latter two cases, we performed 3 pairs of permutation tests (PT vs. NT, PT vs. B, and NT vs. B) and presented corresponding results of US/ES lists as well as SD (specificity diversity) values.

Fig 2 illustrated the distribution of the SS values using the colon cancer as an example: Fig 2A and 2B used the standard histograms, and Fig 2C used the standard box charts. The figure shows that the distributions of the SS values, whether it is of PT or NT samples, are highly skewed, rather than Gaussian-distribution-like symmetrical. It was found that the distribution of species specificity usually follows highly skewed power law distribution and rarely follows bell-shaped (symmetrical) Gaussian distribution (Ma 2023).

**Fig 2.**
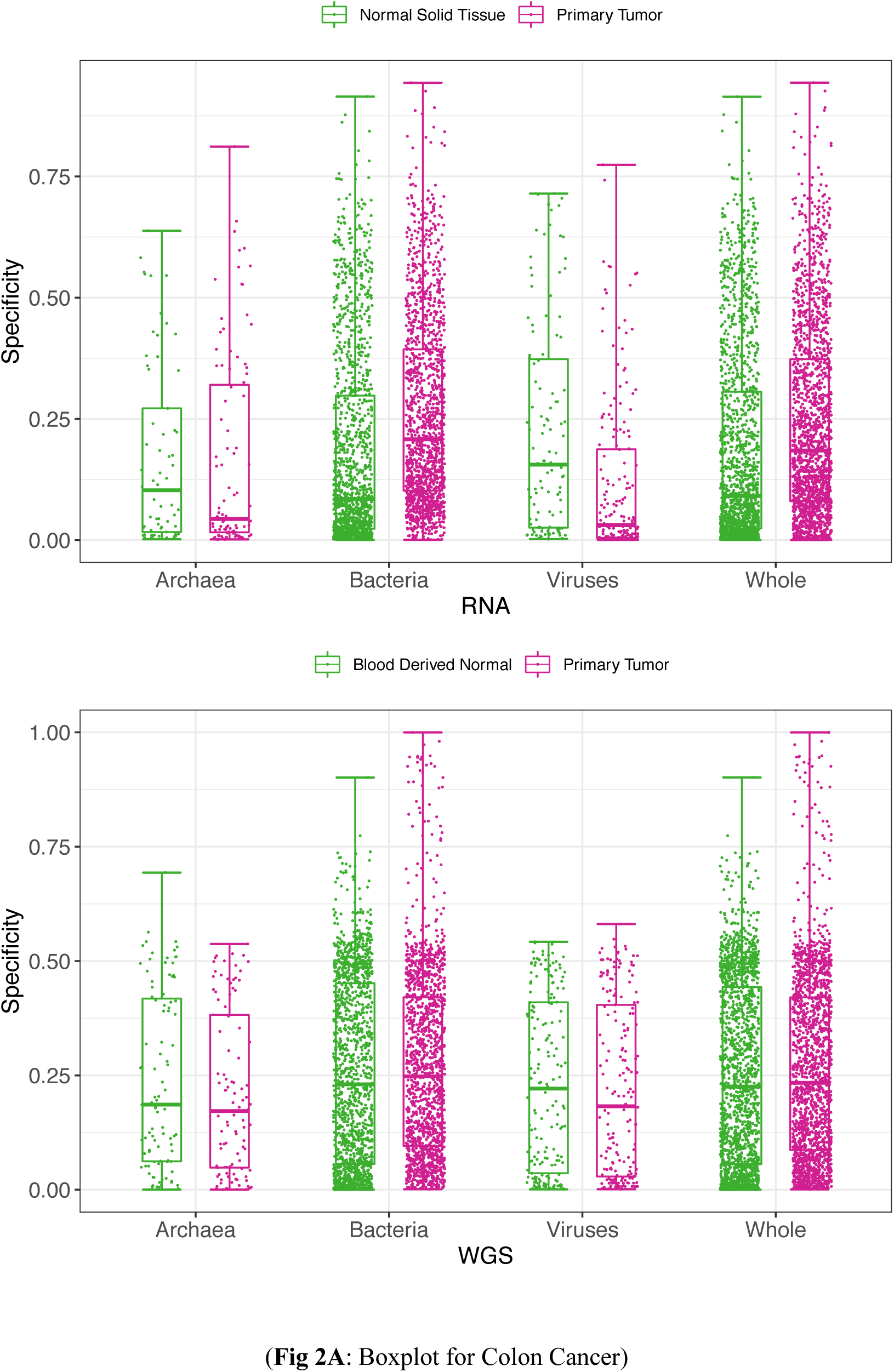
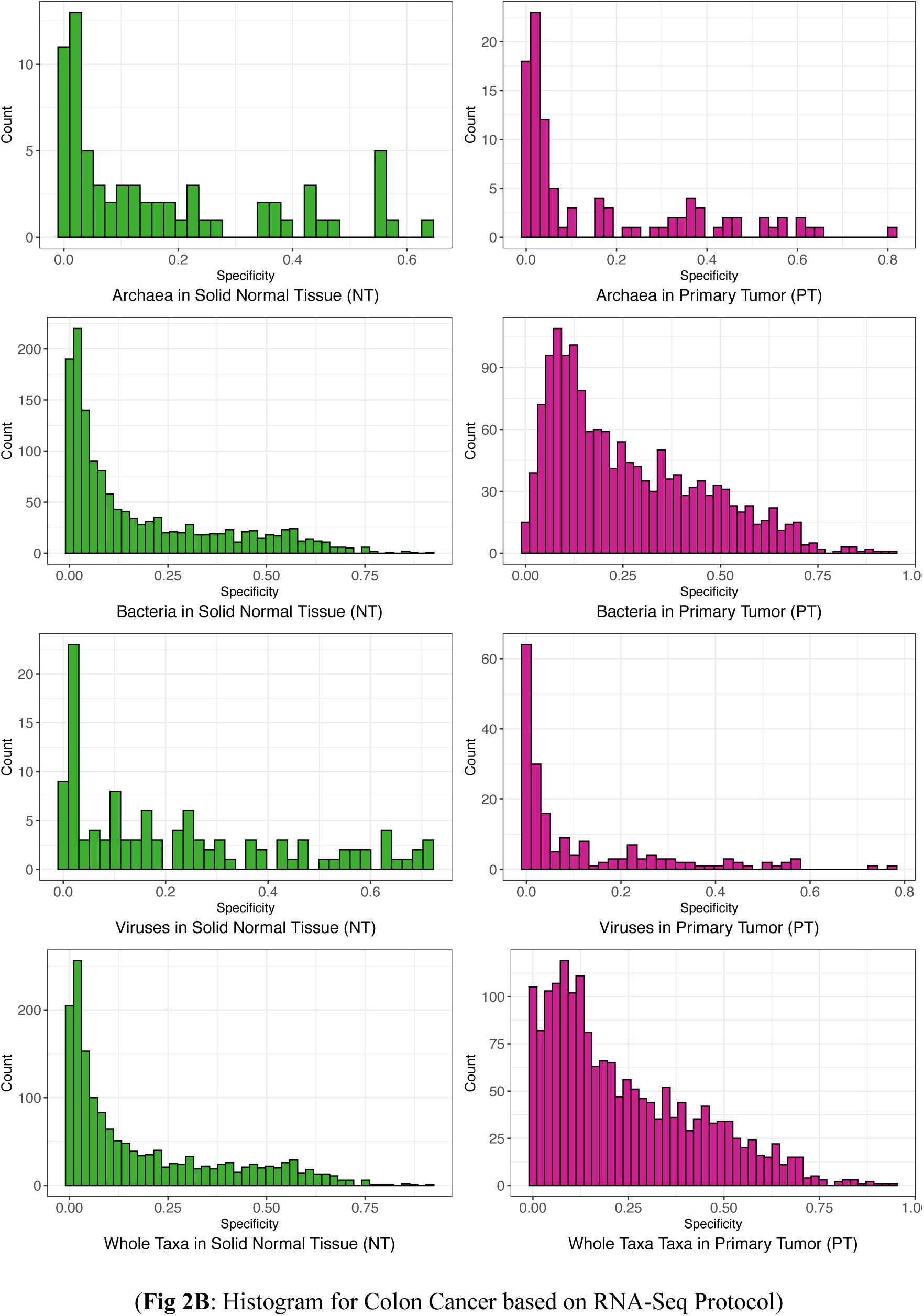
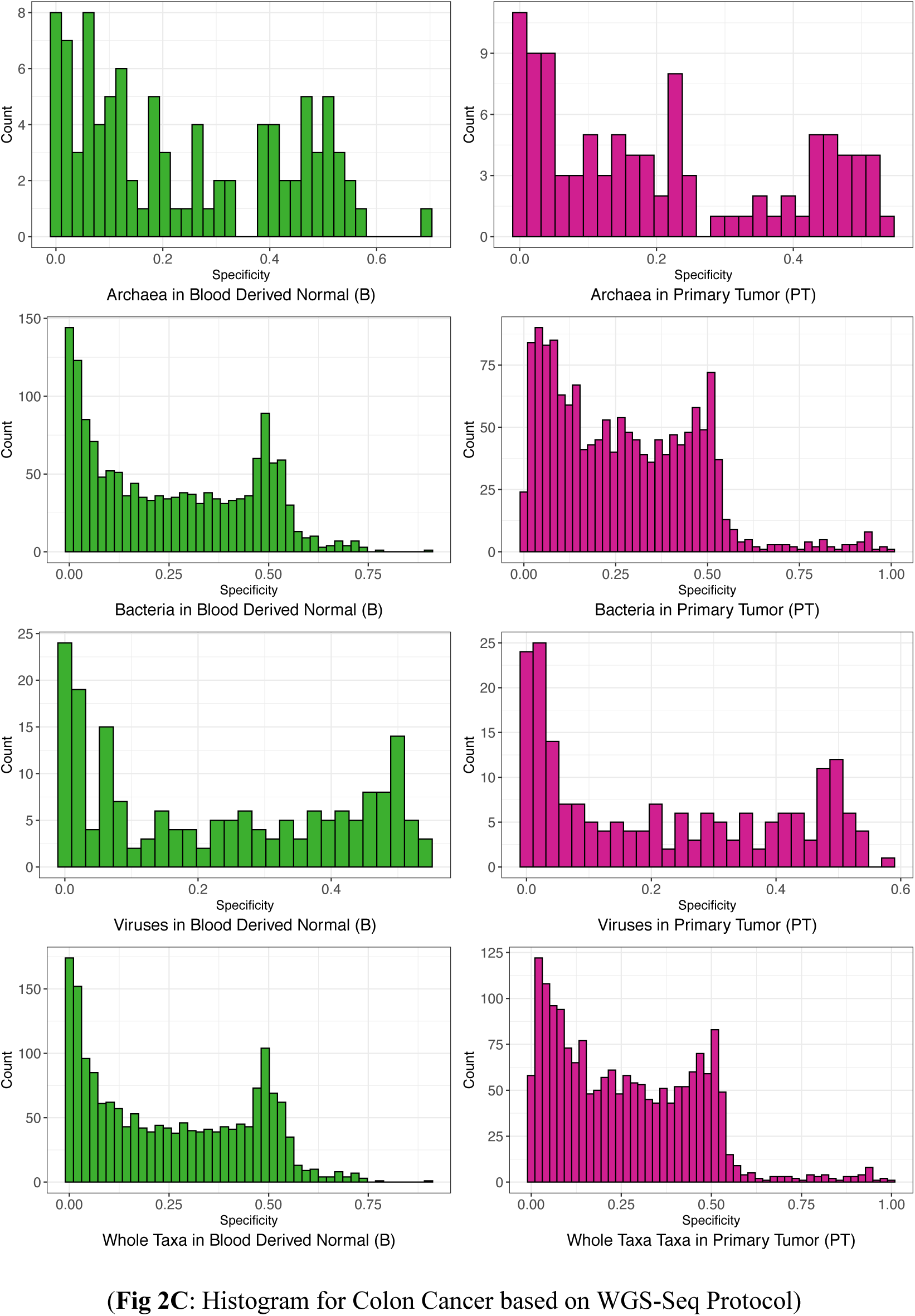
The boxplot (2A) and histograms (2B from RNA-Seq & 2C from WGS) of the frequency distribution of species specificity (SS) values: using the colon cancer as examples.

We then performed species specificity permutation (SP) tests for the treatments (PT, NT/ Blood) of each cancer type (Table S2). Table S2A-S2H (8 sheets) exhibited the SP test results (classifications of species based on SS) for “archaea from RNA-Seq”, “bacteria from RNA-Seq”, “viruses from RNA-Seq”, “whole (all three taxa pooled) from RNA-seq”, “archaea from WGS”, “bacteria from WGS”, “viruses from WGS”, and “whole (all three taxa pooled) from WGS”, corresponding to Table S1A-S1H.

In Table S2, for each combination of the cancer type, taxa (archaea, bacteria, viruses, or whole), and sequencing protocol (RNA-Seq or WGS), the SP test was performed for the combination, and all species in the cancer type are classified as four categories: unique species (US) in PT or NT microbiomes, enriched species (ES) in PT or NT microbiomes, and the ES is equivalent with the impoverished (IS) in the counterpart treatment, *i.e*., ES in the PT equivalent with IS in the NT. Note that, to obtain the four categories of 2 US and 2 ES, we first divide all species into two super-categories: with or without significant differences in species specificity between the PT and NT treatments. For the super-category without significant differences in species specificity, we do not pursue their classification further in this study. For the super-category with significant differences in SS, the SP test is continued to obtain the four categories explained previously. For each species category, the specificity values for each species, the actual difference (delta) in the specificity between NT and PT, the lower and upper bounds of the simulated difference in the specificity between NT and PT from 1000 times of permutation re-sampling, and the *P*-value from the SP test are exhibited (Table S2). When the *P*-value≤0.05 for a species, it is determined that the difference in species specificity (SS) is statistically significant, from which the classification for the species is made.

Fig 3A-3D, again using colon cancer as an example, displayed the “volcano” graphs for the archaea, bacteria, viruses and whole (pooled of the previous three) taxa. In the “volcano” graphs, the Cartesian coordinate system displays five categories (2 USs, 2 ESs, and indifferent species) (*see* the figure legend for the details). An additional advantage of the volcano graph is that it can also distribute (scatter) the species (points) in terms of their relative differences in specificity values.

**Fig 3.**
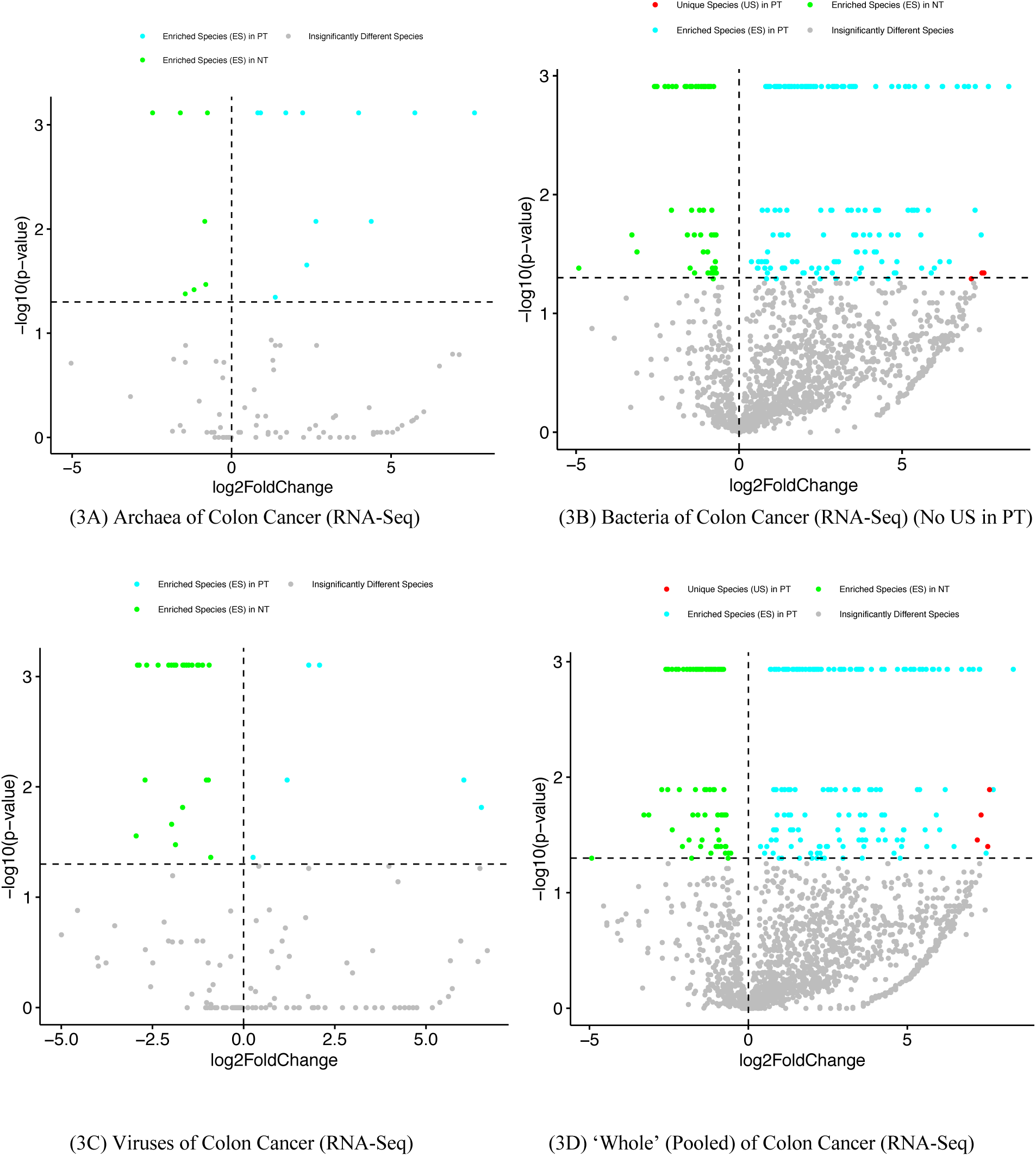
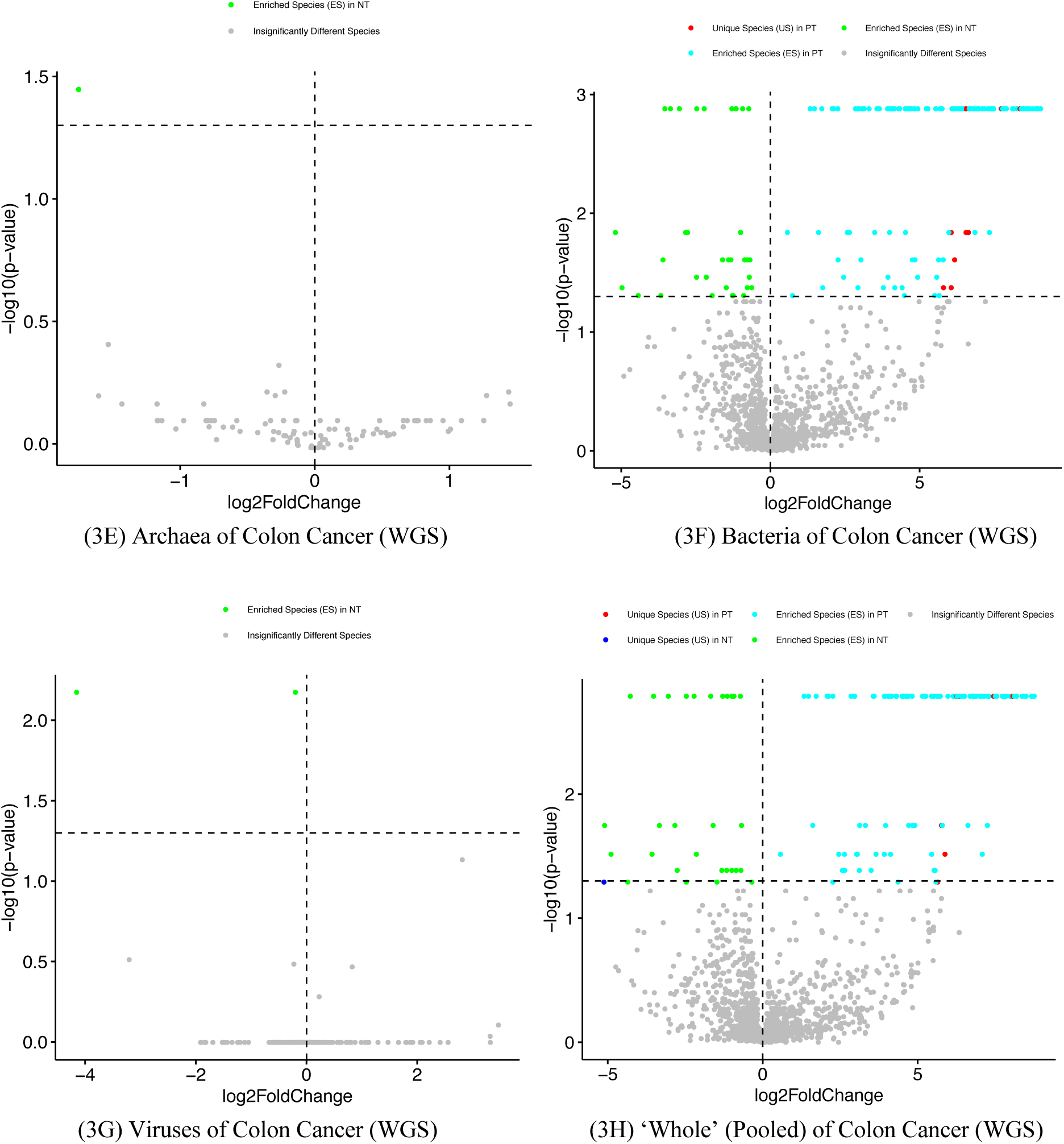
The volcano graphs representing the species classifications for each of the four taxa based on the specificity-permutation algorithm [3A-3D from RNA-Seq: 3A (archaea), 3B (bacteria), 3C (viruses), and 3D (whole=pooled of the previous three); 3E-3H from WGS: 3E (archaea), 3F (bacteria), 3G (viruses), and 3H (whole=pooled of the previous three)]. The *x-*axis represents the log-transformation of the specificity fold change between NT and PT, where fold change=*S_PT_/S_NT_* (*S* represents specificity of species); *y*-axis represents the negative log-transformation of the *P*-value from SP tests of the specificity differences between NT and PT treatments. The vertical dotted line at *x*=0 represents fold change=1 (*i.e*. *S_PT_*=*S_NT_*), the points in the right side of this dotted line represent species with *S_PT_/S_NT_* >1 (*i.e*. *S_PT_*>*S_NT_*), the left points represents species with *S_PT_/S_NT_* <1 (*i.e. S_PT_*<*S_NT_*). The horizontal dotted line represents *P*-value=0.05 [–log10(0.05)=1.301], the points above the line represent species specificity have significant differences between NT and PT, and the points below represent species of non-significant differences in specificity. Therefore, the grey points represent species of non-significant differences in species specificity between healthy (NT or Blood) and diseased (PT), cyan points represent significant enriched species (ES) in diseased (PT), green points represent significant enriched species (ES) in healthy (NT or Blood), red and blue points represent unique species (US) in diseased (PT) and healthy (NT or Blood), respectively. Note that some of the species categories can be missing, for example, Fig 3A lacks USs in both treatments.

### SDP (SD permutation) tests for the holistic differences between species assemblages

After classifying the super-category of all significant species (all microbial species with significant differences in SS between the healthy and diseased treatments) into four different categories (2 USs and 2ESs), plus the super-category itself as fifth category and “all species” as sixth category, we further perform the specificity-diversity permutation (SDP) tests to determine the differences in the SD (specificity diversity) between species categories (also termed assemblages or groups in this article) from different treatments (PT and NT) (Tables S3-S6). The SDP tests are conducted in order to determine the holistic differences between the PT and NT treatments in each of the species categories obtained from the previous SP test.

Fig 4 exhibited the results of the SDP tests for the colon cancer type, including six blocks from top to bottom, corresponding to “US in PT”, “US in NT”, “ES in NT”, “ES in PT”, “All significant species”, and “All species”.

**Fig 4.**
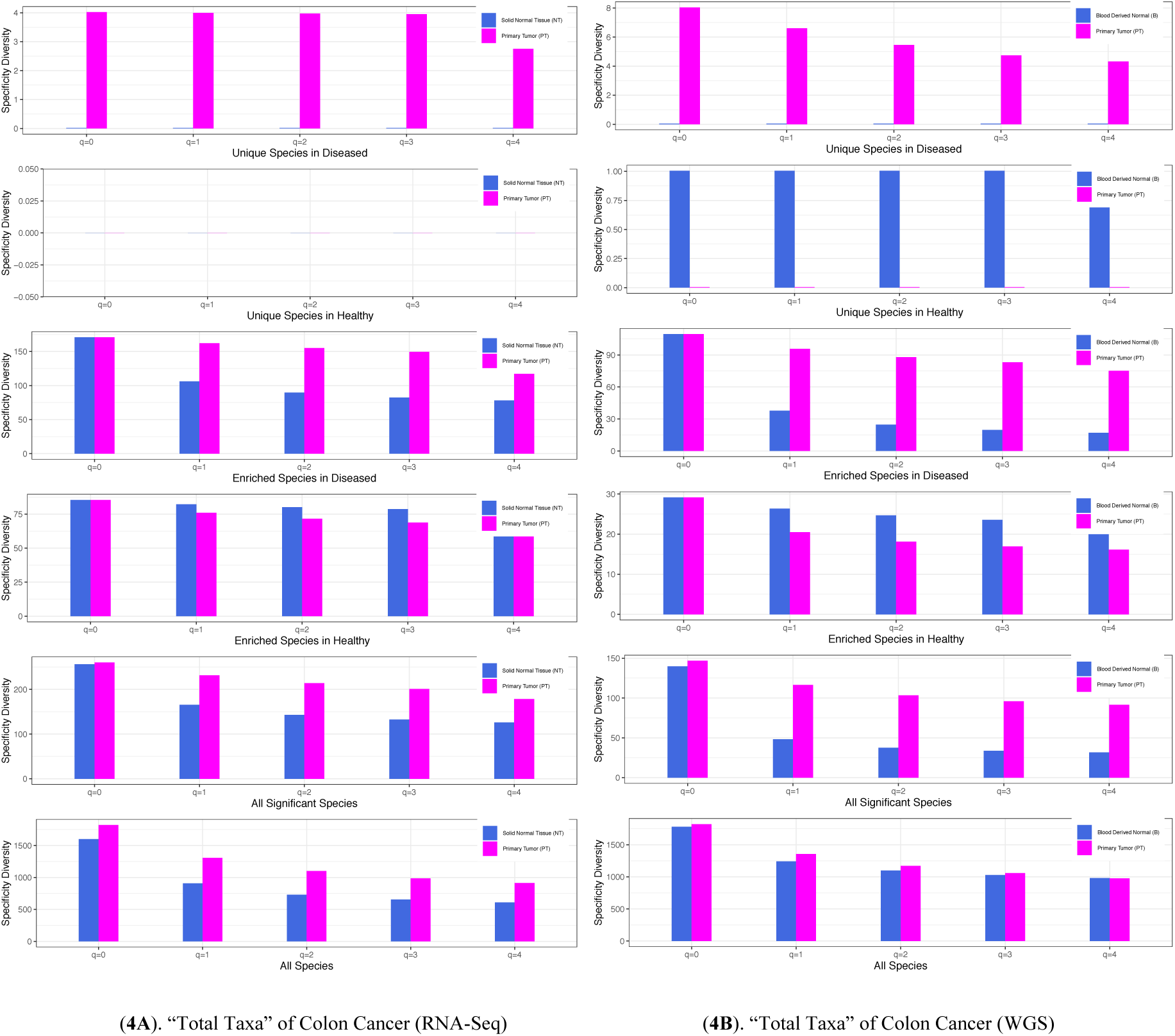
The specificity diversity (SD) for the “total taxa” (pooled of all taxa including archaea, bacteria, and viruses) microbiome datasets of the colon cancer: (4A) Total taxa with RNA-Seq protocol; (4B) Total taxa with WGS protocol. In both (4A) and (4B), from top to bottom, the SDs of unique species (US) in the diseased (PT), unique species (US) in the healthy (NT or Blood), enriched species (ES) in diseased (PT), enriched species (ES) in healthy (NT or Blood), “all significant species”, and “all species” are displayed.

Intuitively, the SD should be significantly different between the NT and PT treatments for the US category (*i.e*., testing the differences of US group in both NT and PT) given the unique (exclusive) nature of the species in either NT or PT (but not in both). In other words, the species in US group only exist in one of the NT or PT exclusively, and their holistic property (SD) should be different, of course. However, as shown by Table S3A and S3B, the SD is significantly different in only 10%-15% cases approximately. The low difference rates are due to the low biomass in tumor tissues, which is likely particularly remarkable for the US species category.

The percentages of differences in SD for the ES species category are modest (55%-63% approximately) as shown in Table S4A and S4B. The differences in SD for the species group of all species with significant differences in SS (species specificity) seems to be the highest (66%-68%) as shown in Table S5. The differences in SD for all species are only approximately 31%-32% as shown in Table S6, which should be expected since the SS values are not in considerations in this case, and the differences should be similar to the differences in species diversity (approximately 1/3, Ma *et al*. 2019). Table S7 further summarized from Table S3-S6 the percentages of difference in SD for various species categories. The SD for specificity diversity order *q*=0 is special in the sense that it is simply the number of species in the species category, for which the SD is computed. It is the same as the species richness in traditional species (community) diversity computation. For this reason, SD (*q*=0) is not included in the previous discussion on the percentages of differences in SD.

**Fig 5.**
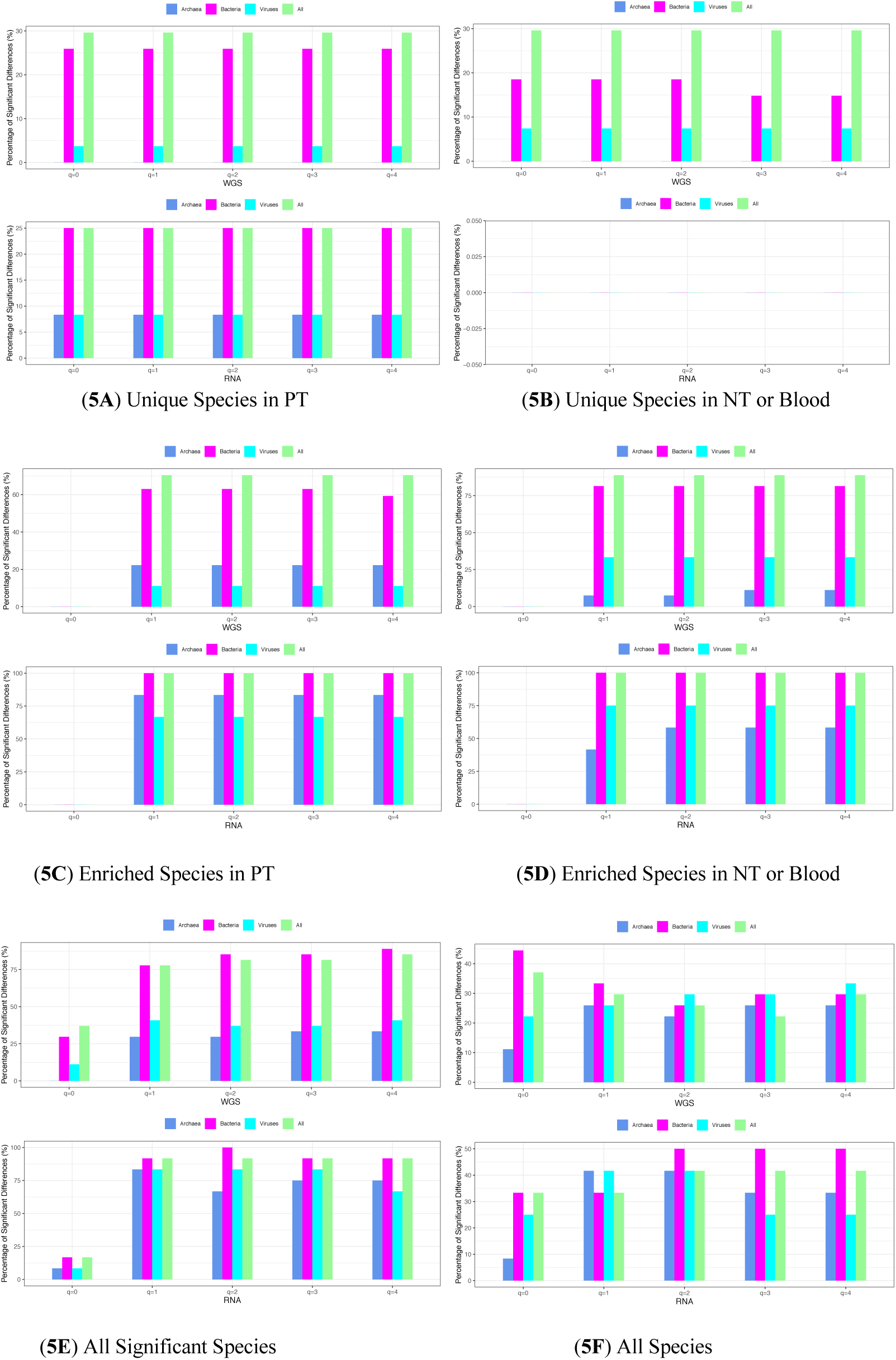
Percentage (%) with significant differences in the SD (specificity diversity) of various species groups between the healthy (NT or Blood) and diseased treatments, computed across all 40x23 possible comparisons of 23 cancer types: The top (5A) & (5B) plots of unique species (US) groups exhibit nearly 100% differences; the middle (5C) & (5D) plots of enriched species (ES) groups exhibit 75%-100% differences; the bottom left (5E) of “all significant species” group exhibits 50%-75% differences; finally the bottom right (5F) of “all species” group exhibits 1/3 differences, all approximately.

### Species specificity network (SSN) and cancer microbiome heterogeneity network (CHN)

We computed the species specificity (SS) lists (tables) for all PT samples, all NT samples, and all Blood samples from the 23 cancer types, respectively, and obtain four species specificity tables (NT & PT from RNA-Seq datasets; Blood and PT from WGS datasets). The computation treats PT (NT, or Blood) of each cancer types as a habitat type. There is a specificity list for each cancer type and all of the lists constitute the species specificity tables (SST), from which we build the SSN and CHN. The Spearman’s correlation coefficients were computed in both manners (as explained below), with FDR-adjusted control (*P*-value=0.01) being used for filtering out statistically insignificant species (nodes/edges) as well as false correlations.

To build species specificity network (SSN), which represents the specificity codependency relationships between species across all cancer types, we compute the pairwise Spearman’s correlation between species in their specificity values across cancer types (*i.e*., for computing the specificity correlation each cancer type is a repetition or case, and the sample size *N*=number of cancer types). Alternatively, to build the cancer microbiome heterogeneity network (CHN), which represents the heterogeneity (homogeneity) between cancer types across all microbial species, we compute the pairwise Spearman’s correlation between cancer types in their species specificity values across species (*i.e*., for computing the specificity correlation, each species is a repetition or case, and the sample size *N*=number of species or OTUs in the species specificity tables).

Fig 6 displays the two SSNs of the PT and NT, built with RNA-Seq datasets (the SSNs of Blood and PT from WGS datasets are omitted to save page space, but included in the following exposition). Some basic network properties are displayed as legends of the network graphs, and those basic properties demonstrated significant differences between PT and NT (with RNA-Seq). It appears that the most significant difference between the primary tumor and healthy tissue (NT or Blood) is that the numbers of network *nodes* in the networks are rather different: for the RNA-seq network, PT=1197 *vs*. NT=264; for WGS, PT=1265 *vs*. Blood=672. Similarly, the edges in PT networks are also much more than those in NT or Blood networks. Since the same statistical criteria [FDR (false discovery rate) control of Spearman’s correlation coefficient with *P*-value=0.01] are used to build the four networks (two are displayed in Fig 6), the disproportionally large number of nodes (species) in the PT networks seem to suggest that the co-dependency between microbial species in PT is increased, compared with that in the NT or Blood. In addition, the network degree (average edges per node) in PT is also larger than that of NT or Blood microbiome networks. Due to these differences, the PT networks are obviously more crowded and densely connected than the NT or Blood networks. We postulate that this phenomenon could be due to potential “stress” or “competition” from tumor cells, which are more demanding in nutrients requirements than the normal cells. This competition may induce or promote close “cooperation” among microbes, as demonstrated by the fact that more nodes and edges passed the FDR control and were admitted into the networks (Fig 6) as explained above. We use the term “cooperation” to represent the rising co-dependency of specificity level in the PT network. Another puzzling change from normal tissue or blood to primary tumor networks is the switch of hub nodes from viruses to bacteria as displayed in Fig 6, for which we do not have an explanation.

**Fig 6.**
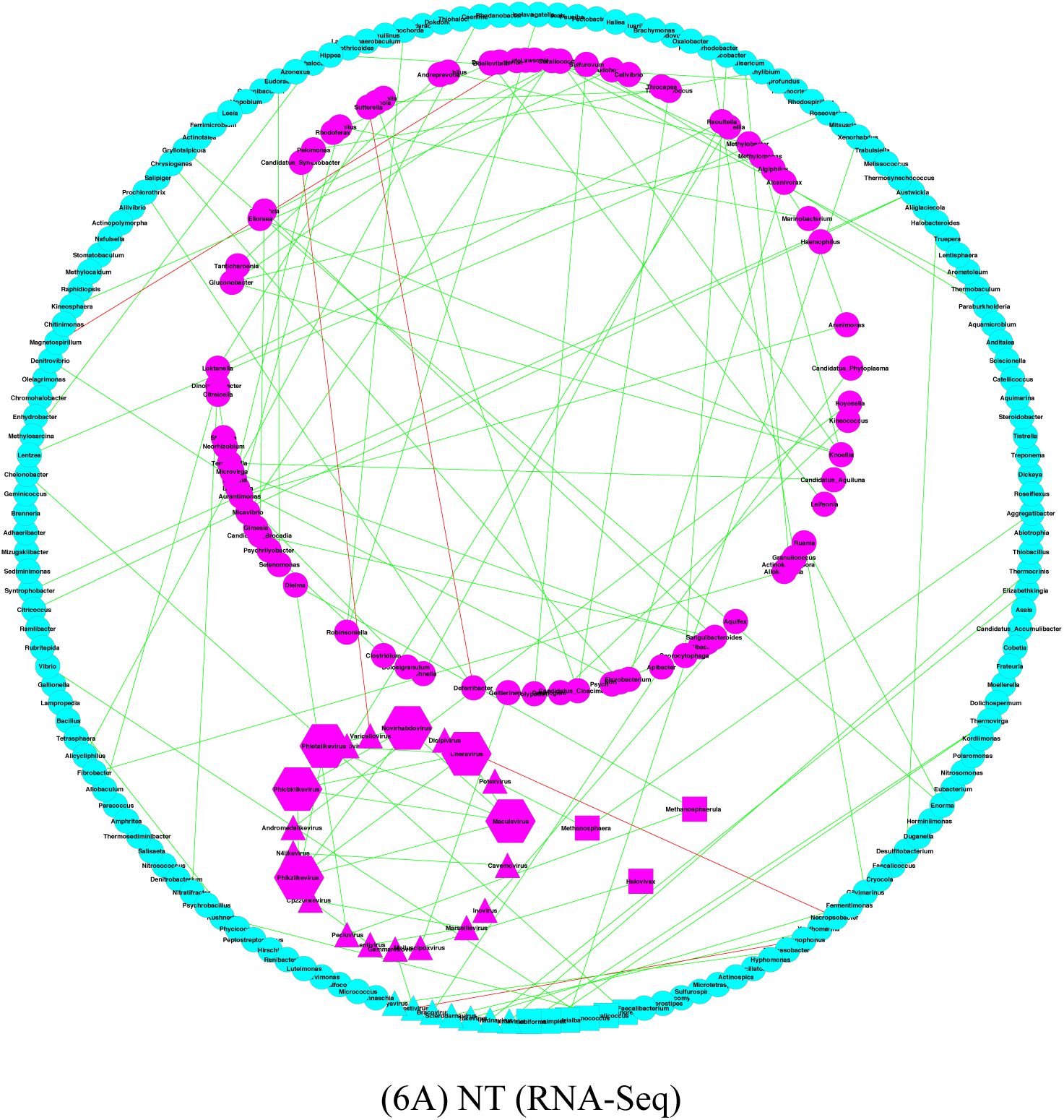
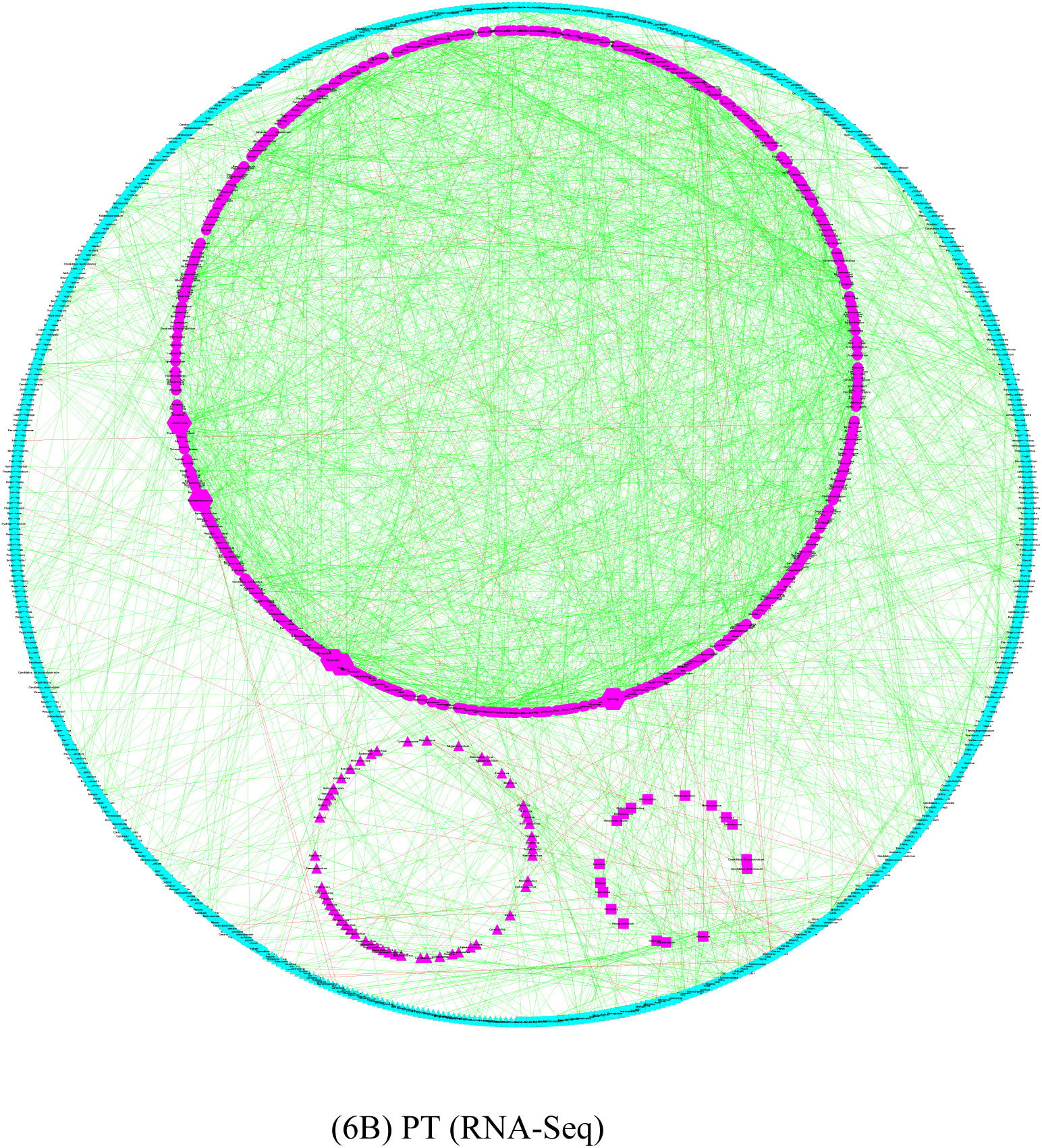
Species specificity network (SSN) for NT (normal tissue) (6A) and PT (primary tumor) (6B) microbiomes, both built with RNA-Seq datasets; legends are as follows, nodes in magenta are core, nodes in cyan are periphery, cycle for bacteria, triangle for virus, square for archaea, hexagons are network hubs, red lines for negative correlations, and green lines for positive correlations. Four microbiomes changes from NT to PT networks: (1) network become more densely connected (more nodes and more edges in PT: 264 nodes & 174 edges in NT, and 1197 nodes & 2498 edges in PT); (2) the average network degree (edges per node) increased from 1.32 to 4.18; (3) the ratio of core to periphery nodes increased from 0.38 to 0.49, *i.e*., more core nodes in PT; (4) hubs are changed from viruses to bacteria. Overall, we postulate that the stress from tumor cells may promote the co-dependency (cooperation) between microbes within tumor microbiomes.

Fig 7 displays the CHN of the PT built with WGS protocol, and the other three networks (Blood for WGS, PT & NT for RNA-Seq) showed the exactly same patterns and are omitted here. All four networks displayed complete graphs (all nodes are connected with each other) for all cancer types (nodes) with sufficient datasets to build the CHN. The message transpired from these networks appears simple: all cancer types appear to show consistent patterns in terms of the specificity co-dependency among microbes. In other words, the microbial codependency appears to be universal across all cancer types, given that all cancer types (nodes) in Fig 7 are connected with each other in terms of their specificity correlations.

**Fig 7.**
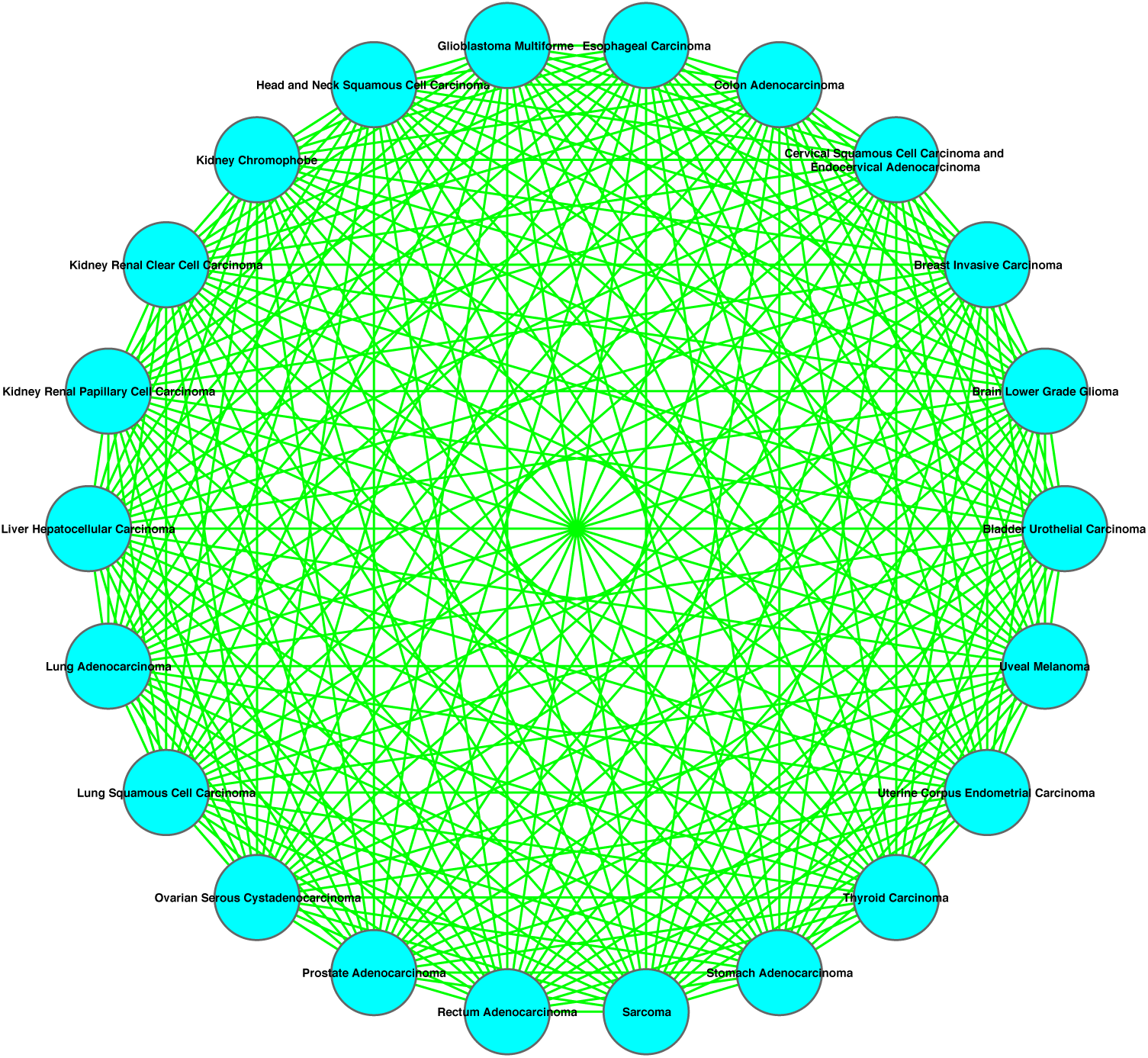
Cancer microbiome heterogeneity network (CHN) for primary tumor (PT) built with WGS datasets: the total graph among 22 cancer types suggests that the previous discussed co-dependency patterns are consistent among cancer types. The CHN for other three treatments (Blood for WGS, NT & PT for RNA-Seq) displayed the same patterns of full graphs.

## Discussion

The significance of this study lies in the successful compiling of the lists of unique and enriched species in each of the 23 cancer types based on specificity permutation (SP) tests, which are of obvious significance for cancer research. Besides those cancer-specific species-level catalogues, we performed assemblage-level tests for the holistic differences based on specificity-diversity permutation (SDP) tests between the healthy (NT or blood) and diseased (PT) treatments for each of the species categories (2 USs, 2 ESs, all significant, and all species). We further conducted specificity network analysis to gain insights on the specificity co-dependency across the landscape of cancer types, which is similar to traditional species co-occurrence network but with a significant difference, *i.e.*, specificity is a global (cross-treatment) property.

Specifically, we made 4 species lists (catalogues) for each of the 23 cancer types (except for two cancer types), including US in PT, US in NT, ES in PT, and ES in NT. In the exceptional two cancer types, which has three treatment (PT, NT and Blood), 6 lists were made for each cancer type from pair-wise permutation tests. In addition, a fifth list of “all species with significant differences” (“all significant species”) is available by combining the previous 4 or 6 lists. Actually, “all species” without considering the specificity differences is considered as a sixth list, which is used in the SDP test. The SDP tests were applied to all six lists (groups) to determine their holistic differences between the PT and NT treatments.

The species specificity of a species is the product of the species’ local (treatment or habitat specific) prevalence (how many samples contain the species in the local treatment) and global (across treatments or habitats) abundance share (fraction of local abundance out of global abundance). When specificity=0 with statistical significance (*P*-value=0.05), the species is determined to be absent in the local habitat (treatment) or unique (exclusive) in its counterpart habitat in the pair-wised SP test. When the specificity value of a species in PT is statistically significantly larger than that in NT, then it is judged to be enriched (up-regulated) in PT based on the pair-wised SP test, which is equivalent to be impoverished in its counterpart (alternative) treatment (NT or blood in this study).

The specificity diversity (SD) is defined for an assemblage (group) of species in terms of Renyi’s (1961) entropy or Hill (1973) numbers at different diversity order (*q*). At diversity order *q*=0, SD defaults to *species richness* (number of species) given that specificity is not weighted in for computing SD (*q*=0). This is the same concept of *species richness* in traditional species diversity in community ecology. At diversity *q*=1, SD is weighed in proportion to the relative specificity (frequency) of each species, and it measures the diversity of species with typical specificity. When diversity *q*>1, SD is weighted in favor for higher specificity species and low specificity species tend to be ignored. The higher diversity order SD tends to represent higher prevalent or abundant species. We used SDP tests to detect the differences in SD between the PT and NT treatments for each of the four species groups (2 ESs, 2USs), the fifth (all significant species) and sixth (all species) groups previously mentioned. The species category of all significantly different species in species specificity (SS) has the highest percentage of SD differences, approximately twice that of the species category of all species (*i.e*., without considering SS), *i.e*., (66%-68%) *vs*. (31%-32%). This contrasting difference highlights the importance of SS in detecting the treatment effects (the tumor effects). For the “all species” group (*i.e*., ignoring SS), the percentage with significant differences is virtually the same as the level exhibited by the traditional species diversity reported by Ma *et al*. (2019). We tend to believe that this same level of 1/3 is actually not coincidental. We postulate that the 1/3-level is a “universal” property when *all species* are included in measuring microbial community properties without resorting to sub-grouping metrics such as species specificity.

The species specificity network (SSN) and cancer microbiome heterogeneity network (CHN) built with species specificity tables prompted us to postulate that the stress or “competition” from cancer cells may promote or induce co-dependency or “cooperation” among cancer tissue microbes as demonstrated by more densely connected networks in PT than in NT. A puzzling phenomenon is that the network hub (the most connected network nodes) is switched from the virus (in NT or Blood) to bacteria in the cancer (PT) microbiome network. We can only speculate that this phenomenon may also be to do with the rising codependency among bacteria cells, given that many viruses in the SSN should be bacteriophages.

Finally, three points including two limitations of our study should be noted. First, the *species specificity* in this article is essentially different from the concept of *specificity* in traditional medical tests (*e.g*., Swift *et al*. 2020), even though both can be related to medical diagnosis. In the latter, the specificity of a test method refers to its capacity in correctly filtering out healthy people (*i.e.,* the true negative rate). Our method, if used for cancer diagnosis, may have its own *specificity*, which is beyond the scope of this article. Second, although we presented the results from both WGS and RNA-Seq datasets, we did not provide comparisons of both sets of the results. There were, at least, two technical difficulties that prevented us from the comparisons: one is that both sequencing protocols are of different targets (all DNA sequences *vs*. transcriptomes), another is the significantly more reads from WGS than those from RNA-Seq (*P*-value<0.05, *see* Poore *et al*. 2020). Although it has been suggested that paired WGS and RNA-Seq of same tissue is more reliable for genomic characterizations of cancers, such practice is still a recent trend (Rusch *et al*. 2018, Werling et al. 2020) and it is likely that majority of the datasets in the TCGA database are from non-paired sequencing operations. These factors make the comparisons of WGS-and RNA-Seq-derived microbiome datasets hardly possible. Despite the limitation, in our opinion, the results presented in this study offer valuable resources and insights for the research community of oncology.

## Data Availability

Availability of datasets
Poore, GD, E Kopylova, Q Zhu, R Knight (2020). Microbiome analyses of blood and tissues suggest cancer diagnostic approach. Nature 579, 567-574. https://doi.org/10.1038/s41586-020-2095-1

## Online Supplementary Information

Table S1. Lists of Species Specificity (SS) Values

Table S2. Lists of Species Groups (US, ES, …) classified with Specificity Permutation (SP) tests

Table S3-S6. Results from Specificity-Diversity (SD) Permutation (SDP) tests

Table S7. The percentage with significant differences in SD (Summarized from Table S3-S6)

## Acknowledgements

This study received funding from the following sources: A National Natural Science Foundation (NSFC) Grant (No. 31970116, 72274192).

## Author contributions

ZSM designed the methodology and analysis scheme, wrote the manuscript. LWL curated and analyzed the data and participated in the results interpretations. JDM conceived the study and participated in the interpretation of the results.

## Ethic Approval

N/A since the study does not involve any wet-lab experiments or survey on human or animal subjects, and all analyzed datasets are already available in public domain, as mentioned above.

## Availability of datasets

Poore, GD, E Kopylova, Q Zhu, … R Knight (2020). Microbiome analyses of blood and tissues suggest cancer diagnostic approach. *Nature* 579, 567–574. https://doi.org/10.1038/s41586-020-2095-1

